# Evidence for immunity to SARS-CoV-2 from epidemiological data series

**DOI:** 10.1101/2020.07.22.20160028

**Authors:** Pablo Yubero, Alvar A. Lavin, Juan F. Poyatos

## Abstract

The duration of immunity to SARS-CoV-2 is uncertain. Delineating immune memory typically requires longitudinal serological studies that track antibody prevalence in the same cohort for an extended time. However, this information is needed in faster timescales. Notably, the dynamics of an epidemic where recovered patients become immune for any period should differ significantly from those of one where the recovered promptly become susceptible. Here, we exploit this difference to provide a reliable protocol that can estimate immunity early in an epidemic. We verify this protocol with synthetic data, discuss its limitations, and then apply it to evaluate human immunity to SARS-CoV-2 in mortality data series from New York City. Our results indicate that New York’s mortality figures are incompatible with immunity lasting anything below 105 or above 211 days (90% CI.), and set an example on how to assess immune memory in emerging pandemics before serological studies can be deployed.

## Introduction

The presence and duration of immunity to novel viruses is traditionally determined through longitudinal serological studies. By characterizing antibodies against a problem virus and tracking the serum levels of these antibodies in a population, for a long enough period, it can be determined with a solid standard of evidence whether the virus induces immunity and how long that immunity lasts.

This method for studying immunity is statistically reliable, but it can demand a very long time and requires ample human and technical resources. Such caveats do not usually pose a problem, but they have become relevant in the case of the recent 2019 coronavirus outbreak. COVID-19 presents the right combination of infectivity and mortality to cause a pandemic of unprecedented global proportions that became clinically and economically relevant in very short timescales, far exceeded by those required for longitudinal serological studies.

Common human coronaviruses causing cold and flulike symptoms of mild degree typically leave an immune memory lasting from six to twenty-eight months [1]. The determinants of coronavirus and rhinovirus immunity have been widely studied for decades and are moderately well understood, as are those of influenza [2]. These diseases leave some immunity, but they can reinfect patients as soon as half a year after.

The mechanisms enabling reinfection strive from simple to elaborate. In the case of influenza and rhinoviruses, highly polymorphic proteins change yearly or faster and thus pathogens escape immune memory through mutation: they are virtually a new pathogen [2, 3]. There is also some evidence that homologous reinfection may contribute to multi-wave influenza outbreaks. Due to previous infections generating an insufficient or non-lasting immune response, recovered patients can become infected again [4]. Other human pathogens such as herpes virus, human cytomegalovirus (HCMV), and human immunodeficiency virus (HIV) elude immunity without fully leaving the human body. This persistence in the face of immune surveillance and medicine is not unique to viruses, as it is well documented in bacteria and tumor cells [5–8]. The populations of these cellular pathogens achieve persistence through the complex interplay of different factors, including extrinsic and intrinsic noise in therapeutic targets, mutation, directly compromising immune function, subpopulations with distinct growth rates, and other phenomena. In viruses, lysogeny often plays a pivotal role, as may do infection of immune cell types.

SARS-CoV-2 is phylogenetically a coronavirus, so the standard of evidence by default would indicate that it induces immunity lasting from one to two years. However, since early in the pandemic, recovered patients have tested positive after previously testing negative. For a while, this rose concerns that SARS-CoV-2 could be not inducing immunity, or persisting in the body after recovery. It is now becoming evident that these positives at least were induced by harmless remains of viral material that endure in the human bloodstream weeks after disease has subsided. But, could immunity after infection be virtually non-existing after all? The presence, extent, and particularities of human immunity to SARS-CoV-2 are still relevant for academics, health professionals and the broader public, and require further research.

Serological studies are the main tool to that aim and continue to unfold as we write this study, with preliminary results already being published [9, 10]. In the meantime, lack of further evidence on human immunity to SARS-CoV-2 delays our full understanding of COVID-19, leads to mismanagement of medical resources such as masks in conditions of scarcity (as the ones we have seen during this pandemic), and sets recovered patients as putative contagion sources, among other undesirable outcomes.

Theoretical alternative approaches to detecting immunity would be desirable in these circumstances; and in principle identifying immunity times should be as simple as inferring the value of a free parameter by fitting an epidemiological model to field data. However, nonlinear dynamical systems like those characteristic of epidemics have a high degree of inherent uncertainty, which makes prediction through conventional, deterministic means inefficient. System variables such as the population of infectious or recovered patients may follow any of a wide array of possible trajectories, and we cannot know which one they will take until they do so [11].

Despite these hurdles, data assimilation techniques are a set of mathematical tools that have provided success in forecasting epidemics [12]. Within these methods, ensemble adjustment Kalman filters (EAKF) have shown capable of providing accurate predictions in a system with many variables [13].

By using Bayes’s theorem to update a model’s predictions with observations at a series of points, uncertainty in a further forecast is reduced, and the span of possible posterior trajectories is limited [14]. The better the measurements (having less uncertainty themselves than the predictions) and the closer in time to the present, the better the updated forecast will be. These Bayesian approaches were originally developed in the context of large-scale geophysical problems and readily and most notably applied to weather predictions [15, 16]. More recently, they have been adapted to epidemiology too, where they became the state of the art in epidemiological forecasting, also in the COVID-19 pandemic, e.g., [17], see also [18] for a related method.

Here, we first examine the impact of immunity memory in the dynamics of a sound epidemiological model of COVID-19. We then estimate the capacity of EAKF techniques to infer the duration of this memory and then apply this approach to mortality time series from New York City (NYC), discerning immunity times against SARS-CoV-2 with reasonable accuracy. Finally, we examine the implications of the presence of immunity in the post-pandemic dynamics. This work thus provides reliable information about human immunity to SARS-CoV-2 and also represents an alternative to longitudinal serological studies for use against future emergent pandemics.

## Results

### Impact of immunity memory on a COVID-19 epidemiological model

We used an epidemiological model in which the total population is divided into a number of classes [19] (Figure 1). The specific compartments represent our current understanding of COVID-19 progression. Note that the infected population is divided into five (right column in Fig. 1) and that we also included two different mortality rates for critical cases because mortality depends strongly on whether there are available beds in Intensive Care Units (ICUs; Methods and Supplementary Material for the model details). All associated parameters in this model are available except for the infection rate *β* and the immunity memory *τ* (Supplementary Material). The fundamental categories resemble those of the well-known SEIRS model [20] in that the recovered population becomes susceptible after some duration of immunity (*τ*). However, the particularities of the COVID-19 progression are such that a minimal SEIRS cannot predict the mid- and long-term dynamics of the population well enough (Fig. S1 and Supplementary Material).

**FIGURE 1.**
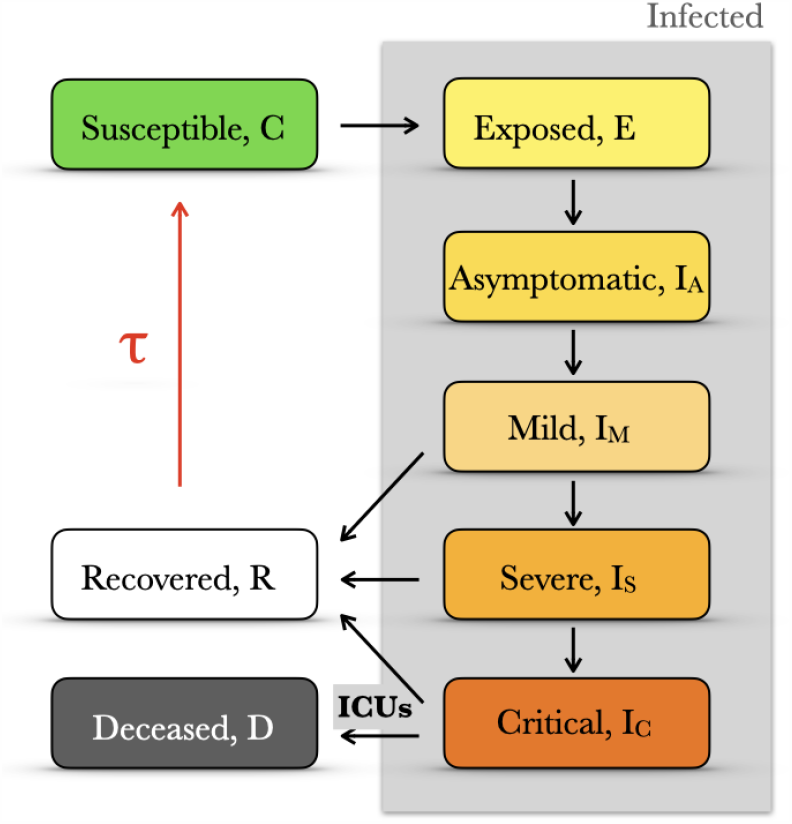
COVID-19 epidemic model. We introduce a compartmentalized epidemiological model with five infected categories (gray shading). Note that the accessibility of ICUs determines the rate to deceased and that those individuals recovered after infection may lose their immunity and become again susceptible after a finite time *τ* (red arrow). See Supplementary Material for model details.

In addition, we performed a variance-based global sensitivity analysis [21, 22] of the model (Fig. S2 and Supplementary Material). In particular, it illustrates the importance of the parameters related to the mild cases in shaping the deceases time series, the peak height and the total number of deceases after a year of pandemic whereas the infection rate tunes mainly the timing of the pandemic peak.

First, we study how the loss of immunity after infection impacts daily deaths (dD/dt) in our COVID-19 epidemiological model. The initial condition is a single exposed case, with a constant and intermediate value of the infection rate (Figure 2A). We find analytically that in the short term, i.e., during the exponential growth of infected cases, the development of immunity has no effect on the initial number of secondary infections *R*_0_ (Supplementary Material): there is not enough time for the re-circulation of the recovered back to the susceptible population.

**FIGURE 2.**
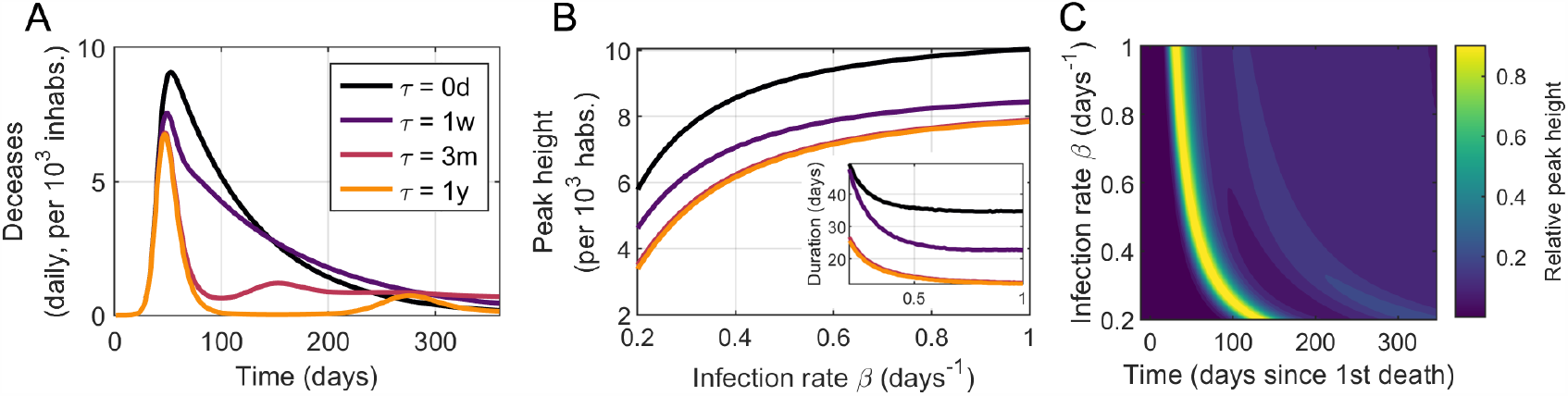
The immunity memory *τ* impacts differently in the mid- and long-term dynamics. (A) Different time series of daily deceases depending on the value of the immunity memory *τ* (*β* = 0.5 days^−1^). (B) The peak height, i.e. maximum number of deceased individuals in a single day, and the duration of the “first-wave” (inset) increase with shorter immunity times. However, the former increases with the infection rate, as opposed to the latter. (C) A finite value of immunity memory induces intrinsic seasonality on the model what produces subsequent epidemic peaks with time. This seasonality depends heavily on the interplay between the infection rate and the immune memory. Data shown for *τ* = 3 months.

However, in the mid-term of the epidemic starting at the departure from exponential growth and up to the first noticeable reduction in daily deceases, a shorter immunity memory time raises the overall number of daily deceases. It also promotes a more prolonged duration of the epidemic as estimated by the time daily deceases stay above 75% of the maximum. Finally, in the long term, beyond the first peak, a finite immunity memory promotes the appearance of new epidemic waves.

The duration of the immunity time also shapes the dependence with *β* of the peak number of daily cases dD/dt_max_ and the duration of the epidemic (Figure 2B). As expected, dD/dt_max_ increases with the infection rate *β* but decreases for increasing *τ*. In addition, we observe that dD/dt_max_ for *τ*s beyond a threshold are hardly distinguishable, e.g., data for *τ* = 3 months and *τ* = 1 year. Moreover, the duration of the first peak decreases together with both increasing *β* and *τ*. Figure 2C displays the intrinsic seasonality derived from a finite immunity time. The height and timing of the secondary peaks are strongly dependent on both *β* and *τ*, data shown is for a fixed value of *τ* = 3 months. This subscribes earlier projections obtained with a multi-strain model [23].

### Predicting immunity memory of an ongoing epidemic

A finite value of the immunity time impacts the time series of daily deceases only after the exponential growth, and starting around the peak of the first wave. To notice these implications, we required a considerably advanced epidemic. What about an ongoing epidemic? This would need the *real-time* assessment of the epidemic parameters and the ability to forecast the short-term dynamics after the epidemic passes the peak.

While this kind of forecasting is intrinsically difficult [13], it is now possible to apply filtering techniques that recently demonstrated valid in this problem by integrating model predictions and data [13, 17, 23, 24]. We thus adopted a specific recursive filtering technique known as EAKF to infer the immunity memory duration in the course of an ongoing epidemic (see Supplementary Material for a brief intro to EAKF) [11, 14].

To describe a typical scenario, we first simulated a *synthetic* time-series with the deterministic model that would represent real data (Figure 3A-C). We then ran 100 independent iterations of the EAKF protocol, with different initial conditions, to estimate dD/dt (everyday deaths) and the “hidden” parameters (*β*_synth_ and *τ*_synth_). To assess the performance of our protocol, we compute the relative errors between the target values and the median of predictions. The similarity between the predicted and real curve of dD/dt is evident (Fig. 3A). Note that the estimates of *β* and *τ* improve mostly before and after the epidemic peak, respectively (Fig. 3B-C). This trend is in agreement with our results from the previous section.

**FIGURE 3.**
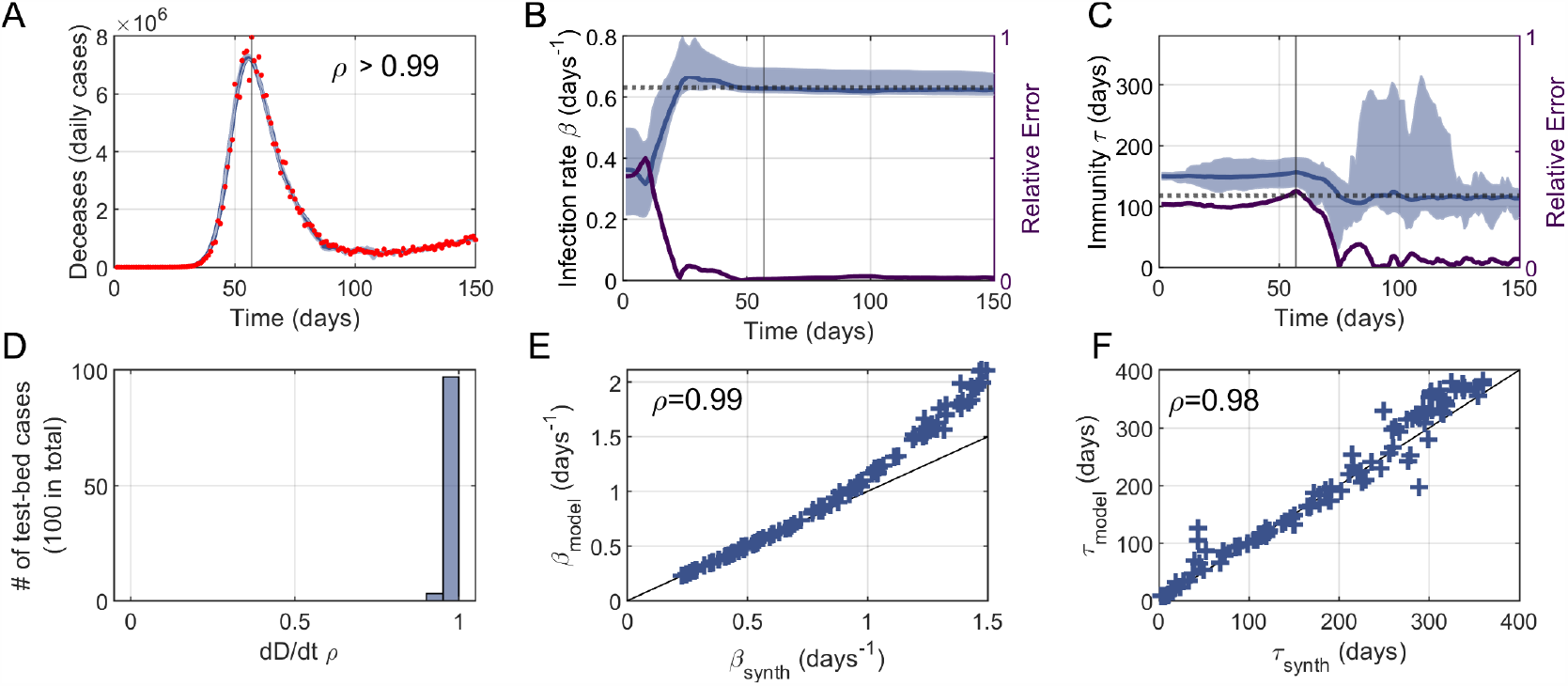
Our EAFK-based approach predicts well the parameters of an ongoing epidemic with synthetic data. We tested the performance of the EAKF-based protocol over a test bed of synthetic data made of 100 time-series generated with the model and random values of *β* and *τ*. Panels A-C illustrate the analysis of a single example whereas panels D-F show the overall performance on the entire test bed. (A) Our protocol (blue) is able to accurately capture data of daily deceases (red), with a linear correlation between data and model *ρ* > 0.99. (B) The value of the synthetic infection rate *β*_synth_ (dotted line) is captured by the protocol *β*_model_ (blue) after some data assimilation steps, and prior to the pandemic peak. Accordingly, the relative error between *β*_synth_ and *β*_model_ decreases as more data is assimilated (purple solid, right y-axis). (C) The immune memory *τ*_model_ (blue) follows similar dynamics as *β*_model_ and approaches the synthetic value *τ*_synth_ (dotted line). However, its relative error (purple solid, right y-axis) drops later than *β*_model_, at about the epidemic’s peak, in agreement with the results of our previous section. In panels A-C, shadings represent 95%CI, while vertical lines denote time of peak. (D) Histogram of linear correlations between model and data of daily deceases (as in panel A) for the entire test bed. (E-F) Synthetic values and EAKF estimates of the infection rate and immune memory largely correlate, *ρ* = 0.99 and *ρ* = 0.98, respectively. We find however that our protocol tends to overestimate larger infection rates when *β*_synth_ > 1.

To further evaluate the limits of this approach, we generated a test bed of 100 synthetic data series for a range of parameters (note that for each series we ran again 100 iterations). Specifically, each series correspond to random values of *β*_synth_ ∈ [0.2, 1.5] days^^−1^^ and *τ*_synth_ ∈ [0, 360] days to which we added relative random noise normally distributed with zero mean and standard deviation up to 10%. The initial exposed population is also selected randomly from a uniform distribution *E*_synth_ (*t* = 0) ∈ [0, 10]. The goal is to apply EAKF within this range to estimate once again the dD/dt series and the “hidden” parameters.

Figure 3D-E shows the performance of our protocol when applied to the entire test bed of synthetic data. On the one hand, we find that infection rates *β* < 1 are excellently captured whereas *β* > 1 are slightly overestimated (Fig. 3E). On the other hand, *τ* is more difficult to estimate in its entire range (smaller correlation between model and synthetic values), but the estimates are not biased towards upper/lower values (Fig. 3F). Moreover we found that the errors between estimates of *τ* and *β* barely correlate (*ρ* = −0.18, not shown), so a better estimation of *β* does not necessarily lead to a worse estimation of *τ*. Also, neither errors in the estimates of *τ* nor *β* correlate with the magnitude of the noise added to dD/dt (*ρ* = 0.04 and *ρ* = −0.12, respectively; not shown).

In sum, filtering and data assimilation techniques successfully identify the values of the infection rate, *β*, and immunity memory, *τ*, when enough data points are available. The value estimates are robustly captured for different initial conditions. Finally, we also found that our protocol can handle up to 10% relative errors with little to no impact on the estimation of *β* and *τ*.

### Quick and strong social distancing measures conceal the mid-term effect of immunity

We now apply the protocol used in the previous section to real time series of new daily deceases reported for COVID-19 in different heavily-affected regions. We performed a preliminary test to rank these regions (world-wide countries and counties/cities within the US) to narrow down potential candidates for signal detection. From over 30 regions, we selected NYC because it had the largest number of deceases per 10^5^ inhabitants and it did not exhibit volatile field data like other regions, e.g. Nassau (NY, USA) or Belgium, (Fig. S3 and S4 in Supplementary Material).

To be certain that the signal in *τ* is not an artifact, we added to the protocol a control variable *δ* that has no effect on the model, it is initialized as a different sample of the same initial distribution as *τ* and follows the same update rules as *τ*. Whether immunity should be considered in the model will depend on its behavior relative to the control variable. The influence of *τ* becomes relevant in the model whenever its distribution deviates significantly from that of *δ*. On the contrary, if immunity mimics the behavior of the control variable it demonstrates that *τ* has no role in shaping the epidemiological dynamics. Thus, statistically significant deviations between the distributions of *τ* and *δ* highlight the influence of *τ* in the results.

In Figure 4A-C we show the results of NYC. The success of the EAKF protocol to capture the dynamics of dD/dt is apparent with a root mean squared error RMSE= 18 deceases and a linear correlation between data and the model median *ρ* > 0.99 (Fig. 4A). In particular, our protocol also captures the time-dynamics of the infection rate, which is well aligned with the days on which NYC promoted social distancing measures: schools and library closings on March 16th, and the pause order of March 22nd (Fig. 4).

**FIGURE 4.**
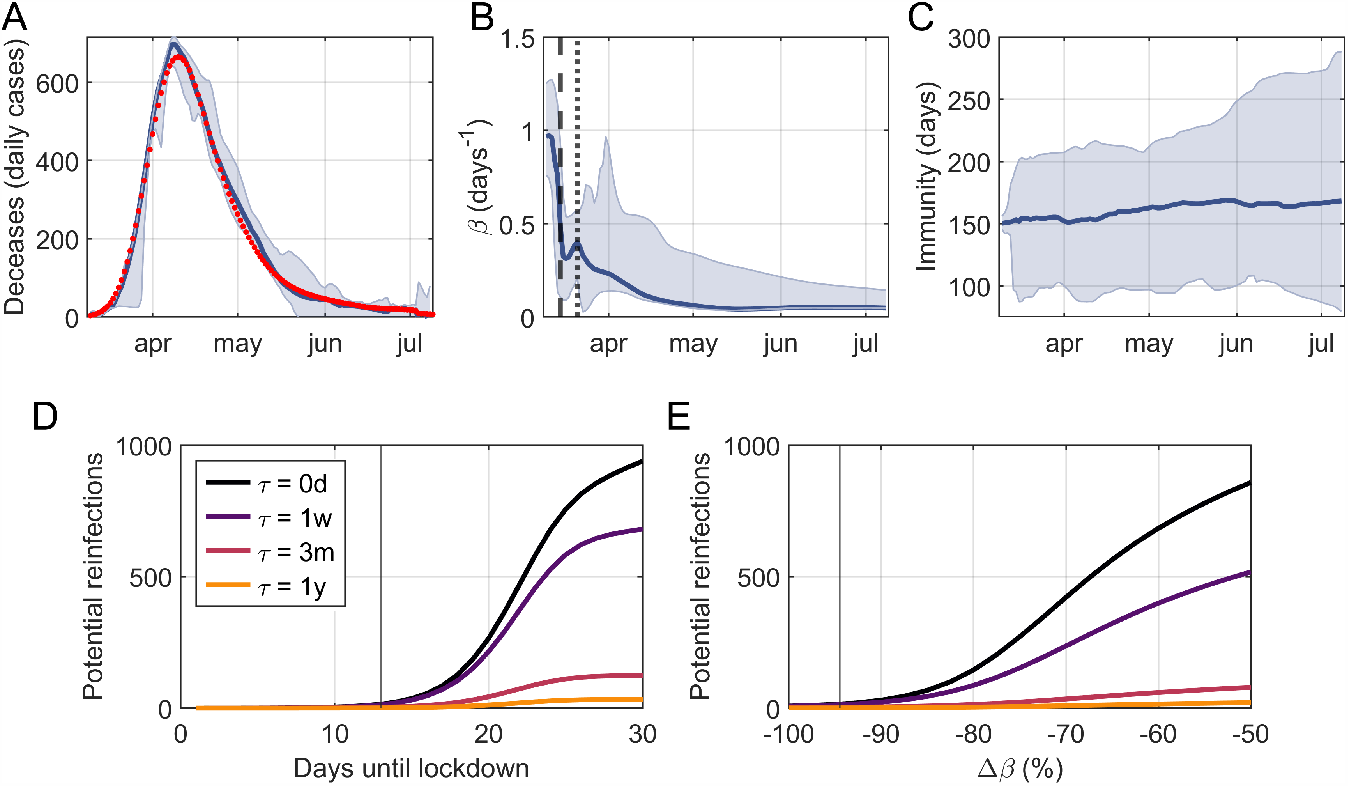
Our protocol captures the days on which social distancing is established, and provide and estimation of immune memory duration with data of New York City (USA). (A) Data (red dots) and algorithm estimate (blue solid, median and 95% CI) of New York City’s daily deceases of COVID-19. Data and prediction are in good agreement, with a root mean squared error RMSE=18 deceases and with a linear correlation coefficient *ρ* => 0.99. Estimate of the infection rate, *β*, dynamics (median and 95% CI). Drops in *β* are well aligned with the days on which social distancing measures took place: school closings (black dashed) and the pause order (black dotted). (C) Estimate of the immune memory duration *τ* (median and 95% CI). The distribution of *τ* becomes significantly different from that of a control variable *δ* (two-sample Kolmogorov-Smirnov test p= 0.017) and sets the lower and upper to *τ* ∈ [80, 288] days with (95% CI). We also simulated a hypothetical scenario based on NYC data with (D) lockdowns established on different days since the first decease, and with (E) different decreases in *β* due to the lockdown. As a proxy for the difficulty of detecting *τ*, we use the number of potential reinfections, i.e. the number of recovered people that has lost its immunity by the 50th day since the start of the epidemic, for different values of *τ*. Observe that specific data for NYC (black vertical lines in panels D and E) illustrates how their quick action in closing schools and passing a pause order (on March 22nd 2020) and their large effectiveness (with *β* decreasing > −90%) ensure a small amount of possible reinfections, and hence the difficulty to capture *τ* with fitting methods.

Most importantly, we found a final estimate of 105 < *τ* < 211 days with 90% confidence (80 < *τ* < 288 days with 95% CI). We obtained this estimate from a statistically significant change in the distribution of *τ* with respect to the control variable *δ* (two-sample Kolmogorov Smirnov, p= 0.017). The upper bound should be considered with caution, given the limited availability of COVID-19 data due to its recent appearance, and future data assimilation steps could alter this bound.

We attribute the difficulty to capture the value of *τ* in real data as opposed to synthetic data to the ubiquity of a strong reduction of the infection rate during the initial days of the epidemic in all data sets that we studied (results of Belgium, Spain and France are available in Fig. S5). We tested this idea by computing the number of recovered cases that have lost their immunity against the virus after 50 days of the start of the epidemic. This number, which we call potential reinfections, is a proxy for the difficulty of capturing *τ*. We consider a scenario similar to NYC, with equal population and equal initial and final infection rates *β*, but with different timing and effectiveness of lockdowns (Fig.4DE). The effectiveness is measured by the relative change between the infection rate pre-and post-lockdown. Simulation data supports our hypothesis since the number of potential reinfections is both close to zero and independent of *τ* when lockdowns are quickly established and/or when they are very effective with infection rate drops > 90%.

Although we confirmed the potential of EAKF algorithms to distinguish the duration of immune memory during an ongoing epidemic, we also noted that the application of our methods is bounded by the expected control measures that are aimed to reduce epidemic progression, i.e., to decrease the infection rate.

### Potential consequences of immunity on post-pandemic COVID-19 dynamics

How could the immune memory for COVID-19 affect a secondary wave of infection? We tackle this by integrating the final state of the EAKF ensembles of NYC with our COVID-19 model deterministically. To account for the relaxation of social distancing measures we include a linear increase of the infection rate during the month of July, specifically, *β* doubles by the 1st of August and remains constant from then on, which is a conservative scenario. In terms of the effective number of secondary infections *R*_*e*_, a doubling of *β* is equivalent to an *R*_*e*_ increase from ∼ 0.7 to ∼ 1.3 (Supplementary Material).

Figure 5 shows the model forecast of daily deceases due to COVID-19 considering the lower and upper bounds of the 95% CI, *τ* = 80 and *τ* = 288 days, respectively. First, in this scenario where social distancing is only slightly relaxed and where the infection rate remains constant from then on, a new epidemic wave in terms of deceases would shortly take place. Its precise timing will depend strongly on the number of ICU beds available, but it can be expected to start in mid September. Moreover, without further social distancing measures during this second epidemic wave, we anticipate that it could last up until January and beyond.

**FIGURE 5.**
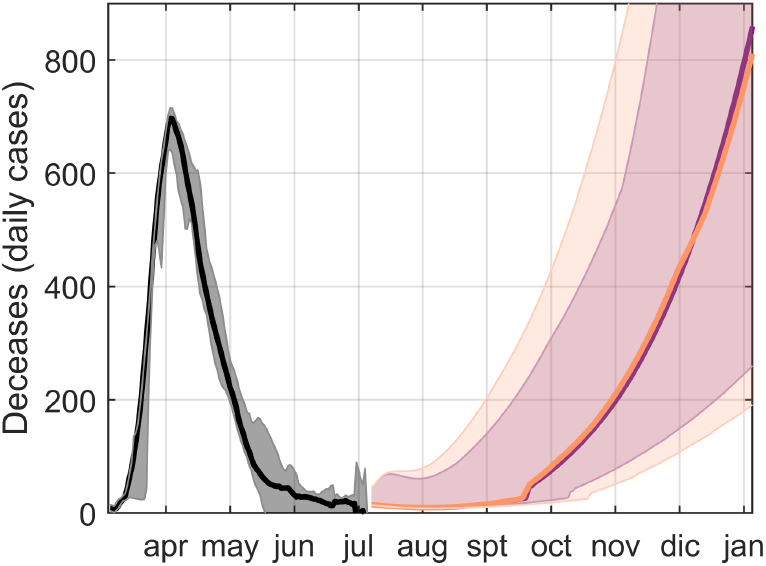
Independently of immunity duration, a second epidemic wave could affect NYC if the infection rate doubles by August. We show the forecast of a second epidemic wave starting in early September if the infection rate *β* doubles its value during July, for two cases of immune memory *τ* = 80 and *τ* = 288 days (purple and orange respectively, shadings denote 95% CI). Observe that *τ* slightly affects the timing of the second wave: the shorter *τ* has a narrower CI. The second wave becomes quickly a real problem due to the little immunity developed during the first (black).

This is due to the fact that, during the first wave, most of the population did not develop immunity to the virus and hence is yet susceptible through the second wave. Such a secondary peak has already been suggested in other specific scenarios [23, 25].

However, if we focus on the maximum effect that different *τ*s have in the short run, we find that, although the median trajectory is independent of *τ*, the confidence region is narrower for *τ* = 80 days. In fact, we expect the immune memory to be relevant only after a considerable fraction of the population has undergone a first infection by COVID-19, or in the case that the time between epidemic waves (or intermittent social distancing) is shorter than that of *τ* where there is enough time to build a sufficiently large pool of immune population.

## Discussion

We propose an alternative approach for estimating the duration of immunity. The protocol relies on the computational analysis of epidemiological time series, which requires far fewer resources and may be deployed faster than its alternatives. Although longitudinal serological studies may be preferred, the evidence for immunity they provide is as indirect as the one we may detect in epidemiological data series. In fact, a direct experimental test of human immunity to SARS-CoV-2 would require intentionally infecting and monitoring recovered human patients with the virus, which would be highly controversial, although this approach has been tested in monkeys [26].

To circumvent this, serological studies obtain indirect evidence based on the premise that antibody prevalence equates immunity, which is generally accurate. However, this is not the case for all diseases. Different mechanisms of persistence deployed by pathogens can uncouple antibody memory from actually being protected against the disease and/or being asymptomatic. Moreover, the effect of immunity on mortality series can hardly be mimicked by any other factors and draws information from field data. Thus, its standard of evidence for immunity is not necessarily lower than the one traditionally employed.

Despite all these points in its favor, the reach of the protocol in its current form is limited, and some requirements must be satisfied to discern immunity. Data series must have surpassed the peak following social distancing measures, which will increase the time necessary to begin a proper examination. In this regard, capturing *τ* was highly dependent on lockdown policies, as evidenced by our potential reinfections metric. The maximum portion of infected people in the population must be sufficiently large for there to be a signal. However, most regions will implement comparable measures to reduce the number of deceased and its growth that make the signal barely distinguishable. In some cases, different stages of social alarm stratified with political or legal restrictions of varying strength are what makes for very volatile infection rates or completely renders changes in immunity irrelevant to early population mortality.

However, segregating exposure and likeliness of infection should improve signal detection as all individuals are not equally likely to be infected. On the one hand, long-lasting cross-immunity with other coronaviruses can significantly reduce the susceptible population [27, 28], and on the other hand, re-infections are most likely occurring in only a subset of the population (such as the working as opposed to the non-working population, age-based classifications, or metropolitan vs suburban or rural). A second way of improving immunity would be to use another observable on top of the deceased during data assimilation. In fact, predictions would improve considerably should data of the infected population be reliable and independent of the limited availability of PCR tests. In addition, the improper mapping of field-measured variables (the “confirmed”, sampling-biased metric) to model variables (exposed, asymptomatic, mild, severe and critical populations) prevents predictability.

But leaving aside reliability in the field tracking of epidemiological variables, it is also worth noting that the protocol is unworkable without a moderately predictive model. Concomitantly, having an accurate model requires some knowledge of the disease’s progression, symptomatology, and outcomes, as well as any notable resources or clinical agents involved in them (as in this case were ICU beds or oxygen). Still, none of these requirements is particularly unlikely to be reached during emergent pandemics. For instance, all of them had been satisfied after 3 months of COVID-19. And the information needed to produce a reasonable model was already public after the second month.

Lastly, perhaps the most significant obstacle in this and more conventional approaches is their inability to discriminate heterogeneity in immunity [29] from groups of recovered patients that have experienced varying degrees of symptomatology. Indeed, patients with many kinds of symptoms and/or peak viral loads may vary in their development of immunity. It could be, for instance, that mild cases do not result in enduring immunity, or result in a shorter immune span, than severe or critical cases. If that were the case, our approach would similarly identify a single overall value for immunity from the statistical overlap of different genuine immunity times, offering a weighed, non-real centrality measure of all immune times in the population.

All these things considered, the present protocol can be thought of as an additional first-hand tool that can always provide necessary evidence in the early stages of a pandemic, until more and varied methods can be deployed.

Now, several issues have arisen surrounding persistence and immunity in COVID-19 throughout the last months. For the majority of the time, the best estimate for immunity to SARS-CoV-2 the community could work with was a presumed range stemming from phylogenetic comparisons pertaining seasonal human coronaviruses like HCoV-OC43 and HCoV-HKU1 [23]. Nevertheless, the standard of evidence of phylogenetic assumptions is not very reliable, particularly with regards to microorganism. According to these suppositions, COVID-19 may elicit immunity lasting from 6 months to 2 years.

Because these were potentially inaccurate measures, early cases of apparent reinfection sparked controversies, and even now as some countries are re-experiencing outbreaks recurring positives are a concern. Our work adds on to other very recent publications that appear to indicate immunity will last at least several months [10]. In particular, a study has observed T cell immunity for SARS-CoV-2 not only in asymptomatic and mild patients, but also in unexposed individuals. This suggests both that the susceptible population has been overestimated and that there are signs of lasting immune memory [30]. In fact, they find similarities with immunity to SARS-CoV-1, pointing that SARS-CoV-2 is likely to have a comparable immune response. Moreover, recent experimental results show that although antibody production depends strongly on disease severity, it lasts at least three months [31] in line with our results. In this context, we provide further evidence that the currently recovered patients will maintain, on average, at the very least 3.5 months of immunity, most likely around 5, and possibly no more than 7; so long as there are no significant differences due to case severity and cross immunity does not provide a far better protection to Sars-Cov-2 than exposition to Sars-Cov-2 itself.

Furthermore, we find that reinfections are a fundamental cornerstone of the current debate on immunity to SARS-CoV-2, especially concerning the development of a vaccine [32, 33]. Although the criteria to label reinfections as such have recently been settled [34], these are demanding. Indeed, to prove a reinfection the patient must have followed a positive-negative-positive PCR testing pattern with at least 28 days between the first and second positives to discard cases of viral remnants [35]. Additionally, it is desirable to have samples of both positives be sequenced and compared to further improve accuracy of re-infection labeling [36]. Also, as in other human coronaviruses, it is highly probable that reinfections produce asymptomatic or mild symptoms [37, 38], in which cases re-testing is hardly expected or not provided by health institutions. However, this might not always be the case, and re-infections can lead to worse symptoms [39]. In general, the probability that a person is symptomatic during both infections, that has been tested negative in between and that samples of both positives are kept and sequenced is rare at best, which highlights the need for surveillance of SARS-CoV-2 reinfections (see also Addendum for further comments on our results).

While we recognize the complexity of the human immune response to SARS-CoV-2, as it is to many other viruses, we trust that our work contributes to a more solid comprehension of the epidemiological implications of this response.

## Materials and Methods

### Data acquisition

We obtained death counts of COVID-19 aggregated by country and USA county from the COVID-19 Data Repository by the Center for Systems Science and Engineering (CSSE) at Johns Hopkins University [40] (last updated on July 9th), and information of ICU beds from a variety of sources depending on the region of study (for the case of NYC see the city’s coronavirus tracker). We used Worldometer to obtain the populations of the regions and countries we selected for analysis [41]. We also used data from the Oxford COVID-19 Government Response Tracker to find the days that different social distancing measures took place in some countries [42].

### Epidemiological model

We introduced a compartmental model that exploits what is currently known about COVID-19 progression and associated accessible data such as the fraction and times at which different infected cases recover or worsen (Supplementary Material, Table S1). Namely the compartments are: susceptible (*S*), exposed to the virus but not yet contagious (*E*), infected and contagious but asymptomatic (*I*_*A*_), with mild symptoms (*I*_*M*_), with severe symptoms (needs hospitalization - *I*_*S*_), and critical symptoms (requires urgent admittance to an ICU - *I*_*C*_); recovered cases (*R*) and the deceased (*D*).

The basic reproductive number, *R*_0_, and its temporal-dependent counterpart *R*_*e*_ (effective reproductive number) are composite parameters that integrate information on not only the infection rate but also the contact rate, susceptible population, and most importantly model architecture [43, 44]. For this reason, we have prioritized the use of the infection rate *β* throughout. However, we have used *R*_*e*_ sparingly due to its biological relevance, which lies in whether it is larger/smaller than the unit, indicating whether the outbreak is expected to continue. To compute *R*_*e*_ we have applied the Next Generation Matrices (NGM) algorithm to our model [45], hence *R*_*e*_ is the largest eigenvalue of the NGM K_*L*_ =-T S^^−1^^ where T and S are respectively the transmissions and transitions matrices (Supplementary Material for more details). A sensitivity analysis of *R*_0_ with respect to the model parameters is available in Fig. S6.

### Data assimilation

The EAKF filtering method consists in propagating and updating ensemble members, which constitute a probabilistic description of the state variables and model parameters [13]. Ensembles are samples of the distributions that the variables are expected to have. In our case, the time-dependent state variables are the infection rate *β*, the immunity memory *τ* and the population in each compartment of the model. We also introduced a dummy variable *δ* that does not affect the model results against which to test the ensemble dynamics of *τ*. The time-dependent observable is the number of daily deceases officially reported, to which we applied a 2 week running average to account for and reporting delays.

In the data assimilation step, the ensemble members are integrated with the model to obtain their expected state at the time of the *succeeding* observation. Next, together with the likelihood distribution of the actual observation, the algorithm calculates the posterior probability assuming that all distributions are normal. Lastly, the unobserved state variables are updated according to their correlation with the observable. For the assimilation of the next data-point, the posterior probability then becomes prior. A more detailed description of the protocol is available in the Supplementary Material.

Importantly, considering that *τ* did not correlate linearly with the observable, we used rank correlations instead to update both *τ* and *δ*. We also used a 3% inflation in the ensemble variance of all variables except *τ* and *δ* since they showed no convergence problems. We have run 100 EAKF instances with ensemble sizes of 200 members. The days in which confinement measures took place (school closing, lockdown…) we added a 200% inflation to better accommodate parameter discontinuities.

## Data Availability

Data availability is discussed in Data acquisition section

https://github.com/CSSEGISandData/COVID-19

## Addendum

### December 17, 2020

After the publication of this manuscript in the medRxiv repository by the end of July 2020, we decided to submit it to several scientific journals for peer review. Some reviewers raised a few issues that unfortunately led to a rejecting editorial consideration even though, in our opinion, they could be argued in their entirety. We take this opportunity to provide a precise answer to these issues further explaining the contributions of this work and also its limitations.

### Rationale and context of this work

By January 2020 both the scientific community and the broader public were already aware of the novel coronavirus SARS-CoV-2, by February it spread without difficulty, and on March 11th the World Health Organization declared the Covid-19 pandemic. During March, most of the countries closed schools, public transport and established a variety of social distancing measures. Initial debates focused on sources of infection (contact surfaces, aerosols), its infectivity (the now well-known *R*_0_), the importance of social distancing, geographical dynamics and, the development of immunity after recovery [46, 47].

While some of these debates have been already clarified, questions around SARS-CoV-2 immunity remain nowadays greatly *unresolved* [48]. In this work, we provided an early estimate of the duration of immunity developed by recovered people after SARS-CoV-2 infections, it being between 3 and 7 months with a 95% CI from New York City (NYC) data. The main concerns raised by readers were

1. the major conclusion is flawed as we obtain a short duration of immunity,
2. this question can only be resolved with approaches characteristic of Molecular Biology,
3. many, many factors determine immunity as to be defined with a single parameter,

which, although already examined in the introduction and the discussion, we will now further review.

### Immunity can be short lasting, and reinfections may be largely underestimated

The principal aim of this work was to explore alternative approaches to assess the duration of immunity given that longitudinal serological studies are time- and resource-consuming. Its main result, which is the duration of immunity, has been questioned for two reasons: immunity appears to last only a few months, and should that be the case, the number of reinfections reported is far less than expected.

Firstly, our estimate of an immune memory lasting several months is in line with initially published results [10]. Moreover, as in the manuscript’s discussion, while we provide a defined range of immunity duration it has to be considered with caution. Partly because not a single range nor a single value can fully describe the high complexity of immunity derived from the ample heterogeneity in viral charges, secondary exposures, disease severity and, individual immune responses [29]. Note that reports on reinfections within a few months after the first infection also support our lower bound to the duration of immunity [39].

Secondly, while some propose that reinfections are a rare event, others argue that most of them are going unnoticed. However, we believe both stances are appropriate as suggests the Covid-19 reinfection tracker [49]. Confirmed reinfections are indeed rare, but the figure grows two orders of magnitude if we also consider suspected reinfections: from 30 *vs*. 1700, respectively. This disparity is partly explained by the lack of genetic proof in suspected cases of reinfections. In addition, most reinfections are hardly detectable as they might present even milder symptoms compared to the first infection.

### Contributions from an alternative approach to the molecular assessment of immunity

The fundamental methodological handicap of our approach is the difficulty with which the duration of immunity is captured during an intense epidemic in which most regions will implement social distancing and confinements as principal contention measures.

In fact, our protocol can only robustly obtain the duration of immunity if reinfections are a significant driver of the epidemic dynamics, as evidenced by the success in treating synthetic data and by our reinfections metric for the case of NYC. Although this poses a major obstacle to an accurate estimation of the duration of immunity for experimental data of the Covid-19, we found that NYC data deviated significantly from a reinfection-free scenario, which led to a *significant* estimation of this parameter.

The estimate we provide relies on a simple compartmentalized model, a common approach in the study of epidemics with its limitations, yet tailored to the typical stages of SARS-CoV-2 infection in which the effects of reinfections produced by immunity loss cannot be mimicked by any other factor. To this model, we applied state-of-the-art probabilistic methods that have already been applied previously with success to highly non-linear systems like weather forecasting, and other epidemics [13, 24]. For this reasons, we believe that although limited by data, we provided a sound initial estimate on immunity duration of comparable value to other approaches after only four months since the declaration of the pandemic.

### Conclusion

In this work, we demonstrate that epidemiological models together with state-of-the-art numerical methods are complementary to traditional approaches in providing estimates of the duration of immunity during the Covid-19 pandemic. Finally, the success in analysing synthetic data highlights the potential of this methodology for epidemiological studies beyond Covid-19. We hope that this and other lessons will contribute to a better response to future pandemics.

## References

1. Edridge, A. W. D. et al. Seasonal coronavirus protective immunity is short-lasting. Nature Medicine. https://doi.org/10.1038/s41591-020-1083-1 (Sept. 2020).

2. Jones, S. et al. Evolutionary, genetic, structural characterization and its functional implications for the influenza A (H1N1) infection outbreak in India from 2009 to 2017. Scientific Reports 9, 14690. issn: 2045-2322. https://doi.org/10.1038/s41598-019-51097-w (Oct. 2019).

3. Palmenberg, A. C. et al. Sequencing and Analyses of All Known Human Rhinovirus Genomes Reveal Structure and Evolution. Science 324, 55–59. issn: 0036-8075. eprint: https://science.sciencemag.org/content/324/5923/55.full.pdf. https://science.sciencemag.org/content/324/5923/55 (2009).

4. Camacho, A. & Cazelles, B. Does homologous reinfection drive multiple-wave influenza outbreaks? Accounting for immunodynamics in epidemiological models. Epidemics 5, 187–196. issn: 1755-4365. https://www.ncbi.nlm.nih.gov/pmc/articles/PMC3863957/. (Dec 2013).

5. Beste, D. J. V. et al. The genetic requirements for fast and slow growth in mycobacteria. eng. PloS one 4, e5349.–e5349. issn: 1932-6203. https://doi.org/10.1371/journal.pone.0005349 (Apr. 2009).

6. Li, Y. & Zhang, Y. PhoU is a persistence switch involved in persister formation and tolerance to multiple antibiotics and stresses in Escherichia coli. eng. Antimicrobial Agents and Chemotherapy 51, 2092–2099. issn: 0066-4804 (June 2007).

7. Sharma, S. V. et al. A chromatin-mediated reversible drug tolerant state in cancer cell subpopulations. Cell 141, 69–80. issn: 0092-8674. https://www.ncbi.nlm.nih.gov/pmc/articles/PMC2851638/ (2020) (Apr. 2010).

8. Cohen, A. A. et al. Dynamic Proteomics of Individual Cancer Cells in Response to a Drug. Science 322, 1511– 1516. issn: 0036-8075. eprint: https://science.sciencemag.org/content/322/5907/1511.full.pdf. https://science.sciencemag.org/content/322/5907/1511 (2008).

9. Bobrovitz, N. et al. Lessons from a rapid systematic review of early SARS-CoV-2 serosurveys. medRxiv. eprint: https://www.medrxiv.org/content/early/2020/05/14/2020.05.10.20097451.full.pdf. https://www.medrxiv.org/content/early/2020/05/14/2020.05.10.20097451 (2020).

10. Seow, J. et al. Longitudinal observation and decline of neutralizing antibody responses in the three months following SARS-CoV-2 infection in humans. Nature Microbiology 5, 1598–1607. issn: 2058-5276. https://doi.org/10.1038/s41564-020-00813-8 (Dec. 2020).

11. Gandon, S., Day, T., Metcalf, C. J. E. & Grenfell, B. T. Forecasting Epidemiological and Evolutionary Dynamics of Infectious Diseases. Trends in Ecology Evolution 31, 776–788. issn: 0169-5347. http://www.sciencedirect.com/science/article/pii/S0169534716301185 (2016).

12. Ionides, E. L., Breto, C. & King, A. A. Inference for non-linear dynamical systems. en. Proceedings of the National Academy of Sciences 103, 18438–18443. issn: 0027-8424, 1091-6490. http://www.pnas.org/cgi/doi/10.1073/pnas.0603181103 (2020) (Dec. 2006).

13. Shaman, J. & Karspeck, A. Forecasting seasonal outbreaks of influenza. en. Proceedings of the National Academy of Sciences 109, 20425–20430. issn: 0027-8424, 1091–6490. http://www.pnas.org/cgi/doi/10.1073/pnas.1208772109 (2020) (Dec. 2012).

14. Evensen, G. Data assimilation: the ensemble Kalman filter 2nd ed. isbn: 978-3-642-03710-8 (Springer, Dordrecht; New York, 2009).

15. Anderson, J. L. An Ensemble Adjustment Kalman Filter for Data Assimilation. Monthly Weather Review 129, 2884–2903. issn: 0027-0644. eprint: https://journals.ametsoc.org/mwr/article-pdf/129/12/2884/4194211/1520-0493(2001)129\_2884\_aeakff\_2\_0\_co\_2.pdf. https://doi.org/10.1175/1520-0493(2001)129%3C2884:AEAKFF%3E2.0.CO;2 (Dec. 2001).

16. Kalnay, E. Atmospheric modeling, data assimilation, and predictability isbn: 978-0-521-79179-3 978-0-521-79629-3 (Cambridge University Press, New York, 2003).

17. Li, R. et al. Substantial undocumented infection facilitates the rapid dissemination of novel coronavirus (SARS-CoV-2). en. Science 368, 489–493. issn: 0036-8075, 1095-9203. https://www.sciencemag.org/lookup/doi/10.1126/science.abb3221 (2020) (May 2020).

18. Dehning, J. et al. Inferring change points in the spread of COVID-19 reveals the effectiveness of interventions. Science 369. issn: 0036-8075. eprint: https://science.sciencemag.org/content/369/6500/eabb9789.full.pdf. https://science.sciencemag.org/content/369/6500/eabb9789 (2020).

19. Kermack, W. O. & McKendrick, A. G. Contributions to the mathematical theory of epidemics–I. 1927. eng. Bulletin of Mathematical Biology 53, 33–55. issn: 0092-8240 (1991).

20. Bjørnstad, O. N., Shea, K., Krzywinski, M. & Altman, N. The SEIRS model for infectious disease dynamics. Nature Methods 17, 557–558. issn: 1548-7105. https://doi.org/10.1038/s41592-020-0856-2 (June 2020).

21. Sobol, I. Global sensitivity indices for nonlinear mathematical models and their Monte Carlo estimates. Mathematics and Computers in Simulation 55. The Second IMACS Seminar on Monte Carlo Methods, 271–280. issn: 0378-4754. http://www.sciencedirect.com/science/article/pii/S0378475400002706 (2001).

22. Saltelli, A. et al. Variance based sensitivity analysis of model output. Design and estimator for the total sensitivity index. Comput. Phys. Commun. 181, 259–270 (2010).

23. Kissler, S. M., Tedijanto, C., Goldstein, E., Grad, Y. H. & Lipsitch, M. Projecting the transmission dynamics of SARS-CoV-2 through the postpandemic period. en. Science 368, 860–868. issn: 0036-8075, 1095-9203. https://www.sciencemag.org/lookup/doi/10.1126/science.abb5793 (2020) (May 2020).

24. Yang, W., Lipsitch, M. & Shaman, J. Inference of seasonal and pandemic influenza transmission dynamics. Proceedings of the National Academy of Sciences 112, 2723–2728. issn: 0027-8424. eprint: https://www.pnas.org/content/112/9/2723.full.pdf. https://www.pnas.org/content/112/9/2723 (2015).

25. Yamana, T., Pei, S., Kandula, S. & Shaman, J. Projection of COVID-19 Cases and Deaths in the US as Individual States Re-open May 4,2020. medRxiv. eprint: https://www.medrxiv.org/content/early/2020/05/13/2020.05.04.20090670.full.pdf. https://www.medrxiv.org/content/early/2020/05/13/2020.05.04.20090670 (2020).

26. Deng, W. et al. Primary exposure to SARS-CoV-2 protects against reinfection in rhesus macaques. Science. issn: 0036-8075. eprint: https://science.sciencemag.org/content/early/2020/07/01/science.abc5343.full.pdf. https://science.sciencemag.org/content/early/2020/07/01/science.abc5343 (2020).

27. Le Bert, N. et al. SARS-CoV-2-specific T cell immunity in cases of COVID-19 and SARS, and uninfected controls. Nature. issn: 1476-4687. https://doi.org/10.1038/s41586-020-2550-z (July 2020).

28. Braun, J. et al. SARS-CoV-2-reactive T cells in healthy donors and patients with COVID-19. Nature. issn: 1476-4687. https://doi.org/10.1038/s41586-020-2598-9 (July 2020).

29. Antia, A. et al. Heterogeneity and longevity of antibody memory to viruses and vaccines. PLOS Biology 16, 1– 15. https://doi.org/10.1371/journal.pbio.2006601 (mAug. 2018).

30. Sekine, T. et al. Robust T cell immunity in convalescent individuals with asymptomatic or mild COVID-19. Cell. issn: 0092-8674 http://www.sciencedirect.com/science/article/pii/S0092867420310084 (2020).

31. Ripperger, T. J. et al. Detection, prevalence, and duration of humoral responses to SARS-CoV-2 under conditions of limited population exposure. medRxiv. eprint: https://www.medrxiv.org/content/early/2020/08/16/2020.08.14.20174490.full.pdf. https://www.medrxiv.org/content/early/2020/08/16/2020.08.14.20174490 (2020).

32. Iwasaki, A. What reinfections mean for COVID-19. The Lancet Infectious Diseases. https://doi.org/10.1016/s1473-3099(20)30783-0 (Oct. 2020).

33. Overbaugh, J. Understanding protection from SARS-CoV-2 by studying reinfection. Nature Medicine. https://doi.org/10.1038/s41591-020-1121-z (Oct. 2020).

34. Tomassini, S. et al. Setting the criteria for SARS-CoV-2 reinfection – six possible cases. Journal of Infection. https://doi.org/10.1016/j.jinf.2020.08.011 (Aug. 2020).

35. Wölfel, R. et al. Virological assessment of hospitalized patients with COVID-2019. Nature 581, 465–469. https://doi.org/10.1038/s41586-020-2196-x (Apr. 2020).

36. To, K. K.-W. et al. Coronavirus Disease 2019 (COVID-19) Re-infection by a Phylogenetically Distinct Severe Acute Respiratory Syndrome Coronavirus 2 Strain Confirmed by Whole Genome Sequencing. Clinical Infectious Diseases. https://doi.org/10.1093/cid/ciaa1275 (Aug. 2020).

37. Nachmias, V., Fusman, R., Mann, S. & Koren, G. The first case of documented Covid-19 reinfection in Israel. ID-Cases 22, e00970. https://doi.org/10.1016/j.idcr.2020.e00970 (2020).

38. Gupta, V. et al. Asymptomatic Reinfection in 2 Health-care Workers From India With Genetically Distinct Severe Acute Respiratory Syndrome Coronavirus 2. Clinical Infectious Diseases. https://doi.org/10.1093/cid/ciaa1451 (Sept. 2020).

39. Tillett, R. L. et al. Genomic evidence for reinfection with SARS-CoV-2: a case study. The Lancet Infectious Diseases. https://doi.org/10.1016/s1473-3099(20)30764-7 (Oct. 2020).

40. COVID-19 Data Repository by the Center for Systems Science and Engineering at Johns Hopkins University https://github.com/CSSEGISandData/COVID-19/tree/master/csse_covid_19_data/csse_covid_19_time_series (2020).

41. Worldometer https://www.worldometers.info/ (2020).

42. Blavatnik School of Government. Oxford COVID-19 Government Response Tracker https://www.bsg.ox.ac.uk/research/publications/variation-government-responses-covid-19 (2020).

43. Delamater, P. L., Street, E. J., Leslie, T. F., Yang, Y. T. & Jacobsen, K. H. Complexity of the Basic Reproduction Number (R 0). Emerging Infectious Diseases 25, 1– 4. issn: 1080-6040, 1080-6059. http://www.nc.cdc.gov/eid/article/25/1/17-1901_article.htm (2020) (Jan. 2019).

44. Linka, K., Peirlinck, M. & Kuhl, E. The reproduction number of COVID-19 and its correlation with public health interventions. medRxiv. eprint: https://www.medrxiv.org/content/early/2020/07/07/2020.05.01.20088047.full.pdf. https://www.medrxiv.org/content/early/2020/07/07/2020.05.01.20088047 (2020).

45. Diekmann, O., Heesterbeek, J. A. P. & Roberts, M. G. The construction of next-generation matrices for compartmental epidemic models. eng. Journal of the Royal Society, Interface 7, 873–885. issn: 1742-5662. https://doi.org/10.1098/rsif.2009.0386 (June 2010).

46. Layne, S. P., Hyman, J. M., Morens, D. M. & Taubenberger, J. K. New coronavirus outbreak: Framing questions for pandemic prevention. Science Translational Medicine 12. issn: 1946-6234. eprint: https://stm.sciencemag.org/content/12/534/eabb1469.full.pdf. https://stm.sciencemag.org/content/12/534/eabb1469 (2020).

47. Altmann, D. M. & Boyton, R. J. SARS-CoV-2 T cell immunity: Specificity, function, durability, and role in protection. Science Immunology 5. eprint: https://immunology.sciencemag.org/content/5/49/eabd6160.full.pdf. https://immunology.sciencemag.org/content/5/49/eabd6160 (2020).

48. Saad-Roy, C. M. et al. Immune life history, vaccination, and the dynamics of SARS-CoV-2 over the next 5 years. Science 370, 811–818. issn: 0036-8075. eprint: https://science.sciencemag.org/content/370/6518/811.full.pdf. https://science.sciencemag.org/content/370/6518/811 (2020).

49. BNO News. COVID-19 reinfection tracker. BreakingNewsOn. https://bnonews.com/index.php/2020/08/covid-19-reinfection-tracker/ (2020).

