## Supplementary Material for "Evidence for immunity to SARS-CoV-2 from epidemiological data series"

July 22, 2020

### 1 COVID-19 epidemiological model

#### 1.1 Model description

We introduced a compartmentalized epidemiological model that we named SEIRSD as it is based on the core elements found in the already known SEIRS models [1] with four population classes: susceptible (S), exposed (E), infected (I), and recovered (R), to which we added a class counting the deceases (D). This model allows recovered cases to become susceptible after some finite time (duration of immune memory) that we denote as $\tau$.

Our model incorporates several infected compartments to account for different known stages of the COVID-19 infection: asymptomatic ($I_A$), with mild symptoms ($I_M$), with severe symptoms that need hospitalization ($I_S$), with critical symptoms that require necessary access to an Intense Care Unit (ICU, $I_C$). Within each of the infected compartments, a fraction of the population $f_{XY}$ worsens and consequently proceeds to the following infection level, while its reciprocal fraction $1 - f_{XY}$ recovers.

The transition between critical cases ($I_C$) and deaths depends strongly on ICU bed availability. To describe this, we calculate the number of ICUs that are occupied by critical cases as $g(I_C; ICU) = min(I_C, ICU)$. All this means COVID-19's main departure from the SEIR archetype (aside from the existence of a contagious asymptomatic stage) is that there are two mortality rates. We discuss the parameters of the model in the next section (see also Table S1). We did not considered ICUs in the test bed to simplify because each country has a different number of ICUs, and because for realistic values of ICUs, $< 10$ per $10^5$ inhabitants, they quickly become irrelevant during an active epidemic as the total number of infected population greatly surpasses that number.

$$\frac{\mathrm{d}S}{\mathrm{d}t} = -\frac{\beta}{N} S \sum_i I_i + \frac{R}{\tau} \tag{1}$$

$$\frac{\mathrm{d}E}{\mathrm{d}t} = \frac{\beta}{N} S \sum_i I_i - \frac{E}{\tau_E} \tag{2}$$

$$\frac{\mathrm{d}I_A}{\mathrm{d}t} = \frac{E}{\tau_E} - \frac{I_A}{\tau_A} \tag{3}$$

$$\frac{\mathrm{d}I_M}{\mathrm{d}t} = \frac{I_A}{\tau_A} - f_{MS}\frac{I_M}{\tau_M} - (1 - f_{MS})\frac{I_M}{\tau_{MR}} \tag{4}$$

$$\frac{\mathrm{d}I_S}{\mathrm{d}t} = f_{MS}\frac{I_M}{\tau_M} - f_{SC}\frac{I_S}{\tau_S} - (1 - f_{SC})\frac{I_S}{\tau_{SR}} \tag{5}$$

$$\frac{\mathrm{d}I_C}{\mathrm{d}t} = f_{SC}\frac{I_S}{\tau_S} - (1 - f_{CD})\frac{g(I_C; ICU)}{\tau_{CR}} - f_{CD}\frac{g(I_C; ICU)}{\tau_{CD}^{ICU}} - \frac{I_C - g(I_C, ICUs)}{\tau_{CD}} \tag{6}$$

$$\frac{\mathrm{d}R}{\mathrm{d}t} = (1 - f_{MS})\frac{I_M}{\tau_{MR}} + (1 - f_{SC})\frac{I_S}{\tau_{SR}} + (1 - f_{CD})\frac{g(I_C; ICU)}{\tau_{CR}} - \frac{R}{\tau} \tag{7}$$

$$\frac{\mathrm{d}D}{\mathrm{d}t} = f_{CD}\frac{g(I_C; ICU)}{\tau_{CD}^{ICU}} + \frac{I_C - g(I_C, ICUs)}{\tau_{CD}} \tag{8}$$

#### 1.2 Model parametrization

Model parameters necessarily came from very recent bibliography by scientific standards, often drawing from *preprint* repositories and otherwise grey literature. However paper methodologies do seem sound and the sources are reliable. When possible, we drew information from meta studies rather than particular institutions or individual papers, e.g., [2].

**There exist two mortality rates.** One of the key elements in most up-to-date models was the conditional mortality rate of patients that get critical symptoms, which depends on whether there are available beds in ICUs. When intensive care is required to keep someone alive, as in 6% of COVID-19 cases (this percentage will change with age distribution and hospital admission across populations), death will occur soon unless the patient in question can be provided with it. Thus, the mortality of COVID-19 will amount to roughly the same number of patients that reach a critical state if ICUs are swamped. If instead, there are ICU beds available, most casualties will be from patients that cannot defeat the disease even when mechanical ventilation, oxygen, dialysis, antibiotics, and other resources are an option. This mortality is far from 100%, and more than half the patients that enter the ICU with COVID-19 leave it recovered [3].

**Mortality in the ICUs.** Different values ranging from 8 to 50% have been reported ever since the pandemic started. Part of this variability could be linked to age distribution and hospital admission in the population considered. Some studies, like those done by the UK's Intensive Care National Audit Research Center [4], received a lot of attention and reported very high mortalities above 5 in every 10 patients. However, these and other studies suffered (to their credit often self-reportedly) of a statistical fallacy: the average time it takes COVID-19 patients to die in the ICU is lower than the time it takes them to recover to the point of not needing intensive care. So studies that follow a population for a fixed number of days rather than to the ultimate outcomes will overestimate mortality. Since we were using New York City (NYC) data, we prioritized information coming from similarly urban, western areas [3]. With this in mind, we settled for $f_{CD} = 0.30$ as a reasonable mortality rate (tests with values up to 0.40 did not considerably affect our results).

**ICU occupancy times.** Different papers have listed different measures of centrality for the time that takes a critical COVID-19 patient to leave the ICU, $\tau_{CD}^{ICU}$, ranging from 4 to 12 days, and for the time it takes them to be discharged should they recover, $\tau_{CR}$, from 6 to 21 days [2]. Here, as in the case of mortality, a plethora of reasons may be responsible for variability, with some of them being biases and artifacts. Some others may stem from genuine population differences owing to physiological, ethological, or political factors. There certainly seemed to be differences due to region of study, with Chinese, Italian, and otherwise western studies apparently forming three separate clusters. However, occupancy for deceased patients was usually lower than occupancy for recovered ones across studies. Taking all this into account, we settled for $\tau_{CD}^{ICU} = 7$ and $\tau_{CR} = 14$ days as reasonable estimates. With these, as with all other parameters, getting an actual reliable functional population average estimate may be impracticable. However, when choosing a value, we do so considering as much evidence as possible, the origin of our data series, and the sensibility of the model to noise in these parameters within arguably accurate ranges. By reasonable, we mean our selection is consistent with data, and they do not significantly favor nor compromise the accuracy of predictions or the estimation of immunity when compared to other values.

**Additional parameters.** The variability within reported ranges in additional parameters (namely, those about the timeline of disease progression and the times to transition from one variable to the next) was less relevant to the model's predictivity than that in ICU-related parameters. Medians and averages for the total time from contagion to the onset of symptoms change from 5 to 7 days, depending on the study, with the majority of the distribution being included between 2 and 14 days [5–7]. The time from contagion to infectivity, or latency time $\tau_E$, is given as 2 or 3 days in the literature [7]. We thus used 2 days for  and 4 days for the time $\tau_A$ that it then takes to show symptoms ($\tau_E + \tau_A = 6$). Chinese/WHO estimations for the remaining progression times [5, 6] and fractions such as the times $\tau_{MR}$ that it takes for mild cases to recover, the time $\tau_{MS}$ that it takes them to report to

**Table S1.** List of parameters and their values references used throughout.

| Parameter | Value |
|---|---|
| Transition times (days) | |
| $\tau_E$ | 2 [7] |
| $\tau_A$ | 4 [5, 7] |
| $\tau_M$ | 7 [5] |
| $\tau_{MR}$ | 8 [5] |
| $\tau_S$ | 1.5 [5] |
| $\tau_{SR}$ | 14 [5] |
| $\tau_{CD}$ | 1/48 |
| $\tau_{CD}^{ICU}$ | 7 [2] |
| $\tau_{CR}$ | 14 [2] |
| $\tau$ | $[80, 288]$ (95% CI) |
| Transition fractions | |
| $f_{MS}$ | 19% [6] |
| $f_{SC}$ | 31% [5] |
| $f_{CD}$ | 30% [3] |

the hospital and/or progress to a severe state or the fraction of total cases that are mild have remained relevant in the face of western epidemics.

#### 1.3 Equivalence with a SEIRS model

To test the equivalence between our model and a minimal -SEIRS model, we computed a set of "equivalent parameters" that would result in equal epidemic exponential growth rates $\mu$ [8]. Starting from our COVID-19 model-specific parameters $\vec{\theta}_{\text{COV}-19} = (\vec{\tau}, \vec{f})$, we obtain $\mu$ and the population distribution when $E_0 = 1$ as a solution to the linearization of the ODE system. Then, we linearize the ODE system of a SEIRS model,

$$\frac{\mathrm{d}E}{\mathrm{d}t} = \beta I - \gamma E \tag{9}$$

$$\frac{\mathrm{d}I}{\mathrm{d}t} = \gamma E - \sigma I \tag{10}$$

$$\frac{\mathrm{d}R}{\mathrm{d}t} = \sigma I. \tag{11}$$

to find the equivalent set of SEIRS parameters $\vec{\theta}_{\text{SEIRS}} = (\gamma, \sigma)$:

$$\gamma = \beta I_0/E_0 - \mu, \text{ and } \sigma = \beta - \mu(1 + E_0/I_0), \tag{12}$$

where $I_0$ is the number of infectious cases when $E_0 = 1$ in both models.

For example, the results are, in the case where the infection rates $\beta_{\text{COV}-19} = \beta_{\text{SEIRS}} = 1$, the total populations $N_{\text{COV}-19} = N_{SEIRS} = 10^6$, the immune memories $\tau_{\text{COV}-19} = \tau_{\text{SEIRS}} = 10^{10}$ and without ICU beds:

$$\lambda = 0.479 \ , \ I_0 = 0.979 \ , \ \gamma = 0.5 \text{ and } \sigma = 0.032. \tag{13}$$

We used numerical calculations because the analytical computation of $\mu$ becomes intricate for complex models [8]. Figure S1 displays the matching dynamics of a SEIRS and our COVID-19 model that yield the exact same initial epidemic dynamics. Observe that although the starting of the epidemic is equivalent, the dynamics at the end of the exponential regime are notably different for the infected and recovered classes.

##### 1.4 Computing $R_e(t)$ with Next Generation Matrices

To obtain the characteristic reproductive numbers for our compartmentalized model, we computed the corresponding next generation matrices following [9]. The transmission and transition matrices, $T$ and $\Sigma$, respectively, reads as:

$$T(t) = \frac{S(t)\beta(t)}{N}\begin{bmatrix} 0 & 1 & 1 & 1 & 1 \\ 0 & 0 & 0 & 0 & 0 \\ 0 & 0 & 0 & 0 & 0 \\ 0 & 0 & 0 & 0 & 0 \\ 0 & 0 & 0 & 0 & 0 \end{bmatrix} \quad \text{and} \quad \Sigma = \begin{bmatrix} \frac{-1}{\tau_E} & 0 & 0 & 0 & 0 \\ \frac{1}{\tau_E} & \frac{-1}{\tau_A} & 0 & 0 & 0 \\ 0 & \frac{1}{\tau_A} & \frac{f_{MS}-1}{\tau_{MR}} - \frac{f_{MS}}{\tau_M} & 0 & 0 \\ 0 & 0 & \frac{f_{MS}}{\tau_M} & \frac{f_{SC}-1}{\tau_{SR}} - \frac{f_{SC}}{\tau_S} & 0 \\ 0 & 0 & 0 & \frac{f_{SC}}{\tau_S} & \frac{-1}{\tau_{CD}} \end{bmatrix} \tag{14}$$

The time-dependent number of secondary infections $R_e(t)$ is computed as the largest eigenvalue of the large-domain NGM matrix $K_L(t) = T(t)\Sigma^{-1}$. Analytically:

$$R_e(t) = \frac{S(t)\beta(t)}{N}\left(\tau_A + \frac{\tau_M \tau_{MR}}{C_1} + \frac{f_{MS}\tau_{MR}\tau_S\tau_{SR}}{C_1 C_2} + \frac{f_{MS} f_{SC} \tau_{CD}^{ICU} \tau_{CR}\tau_{MR}\tau_{SR}}{C_1 C_2 C_3}\right) \tag{15}$$

where $C_1 = \tau_M - f_{MS}\tau_M + f_{MS}\tau_{MR}$, $C_2 = \tau_S - f_{SC}\tau_S + f_{SC}\tau_{SR}$ and $C_3 = \tau_{CD}^{ICU} - f_{CD}\tau_{CD}^{ICU} + f_{CD}\tau_{CR}$.

Observe that in the early dynamics, when $S(t)/N \sim 1$ then $R_e(t)$ is independent of $\tau$. However, in the later dynamics the effect of $\tau$ is reflected on the behaviour of $S(t)$. Therefore, the immune memory $\tau$ has no impact whatsoever in the linear regime of the ODE system, i.e., the initial dynamics of the epidemic.

##### 1.5 Parameter sensitivity of $R_e(t)$

We analyzed the sensitivity of $R_0$ to every other model parameter with its analytical solution computed before (and the parameters as in Table S1). In Fig.S5 we show the local derivative of $R_0$ around different values of $\beta$. We find that $R_0$ depends strongly on $\beta$ in all situations. Surprisingly we also find that the fraction of mild cases that become severe, $f_{MS}$, can be as relevant as $\beta$ in reducing the number of secondary infections of a single individual. These results endorse the two main intervention measures most commonly found across regions: i) social distancing reduces the contact and infection rates, and consequently $\beta$, and ii) expanding hospital resources reduces conceivably $f_{MS}$, the fraction of mild cases that worsen.

#### 2 Introduction to Ensemble Adjustment Kalman Filter algorithms.

##### 2.1 Motivation

Ensemble adjustment Kalman filter (EAKF) methods are a suite of well-known algorithms for data assimilation, first designed for linear models but also helpful for non-linear ones. Despite the many books and online resources, the use (and perhaps misuse) of technical jargon obscures its understanding at first. Our goal here is to present a brief introduction to the uninitiated. See, for instance, [10, 11] for more details and technical references.

#### 2.2 Bayes' theorem in data assimilation

The fundamental component of these methods is the "assimilation" of available data with the model prediction. In these algorithms, data assimilation is based on Bayes' theorem,

$$P(B|A) = \frac{P(A|B)P(B)}{P(A)}, \tag{16}$$

where $P(X|Y)$ is the conditional probability of event $X$ given $Y$, and $P(X)$ is the marginal probability of $X$. In our context, the posterior probability of a new observation B given a series of previous observations $A$, i.e., $P(B|A)$, is proportional to the product of the prior distribution $P(A|B)$, and the likelihood, or marginal probability, of observation B $P(B)$. Here $P(A)$ is considered to be a normalization constant so that the total probability is equal to 1.

After a new data point is assimilated and we have computed the posterior probability, we can then consider it our initial state to assimilate the next data point, e.g., C,

$$P(C|A,B) = \frac{P(A,B|C)P(C)}{P(A,B)}. \tag{17}$$

We can then apply this iteratively to advance in space and/or time and improve our subsequent predictions. To ease the implementation of this approach, we may consider all probability distributions to be normal. In this way, if the prior and the likelihood are normal, the posterior is also normal and its mean and variance can be easily computed as is explained later.

#### 2.3 Algorithm components and definitions

These algorithms have the following minimal ingredients:

- State vectors $\vec{x}(t)$. They describe the system state at a specific time and they are comprised of the state variables and the free parameters, which in these algorithms are necessarily time-dependent.
- The model $\mathcal{M}$. The model allows to obtain the system state in the next time point: $\vec{x}(t) = \mathcal{M}\vec{x}(t-1)$.
- An observable operator $\mathcal{H}$. It turns a model state vector into an observable value: $\vec{y}(t) = \mathcal{H}\vec{x}(t)$. This is because in some cases an observable is not directly a state variable, but rather a function of the state vector.
- Experimental measurements of the observable and their corresponding error: $\mu_\omega(t)$ and $\sigma_\omega(t)$.
- An initial guess for all variables and parameters. During the algorithm initialization, the j-eth variable of the i-eth ensemble member $\vec{x}_{i,j}(t=0)$ is drawn from a random, normal distribution centered around the initial guesses. Although state vectors tend to converge across time, a reasonable initial guess improves the overall performance of the algorithms. Moreover, small variances in the initial distribution are likely to keep the protocol in its linear regime.

#### 2.4 Main algorithm

Once these ingredients are clear, and assuming that we have an observation every $\Delta t = 1$ time units, the algorithm goes as follows:

0. Initialization. Create an ensemble $\epsilon$ of S members. Each member is a state vector with its values drawn from random normal distributions: $\epsilon(0) = \{\vec{x}_i(t=0)\}_{i=1,\ldots,S}$.

1. Integrate model. Integrate every ensemble member to obtain the prior ensemble state at the time of next data point $\tilde{\epsilon}(1) = \{\vec{\tilde{x}}_i(t=0)\}_{i=1,...,S}$.
2. Obtain the prior distribution of your observable. From every ensemble member, obtain your sample of prior observations $\tilde{y}_i(1)$ with $i = 1, ..., S$. From the sample, compute its mean and variance $\mu_{\tilde{y}}(1)$ and $\sigma_{\tilde{y}}(1)$.
3. Compute posterior distribution of observations. Following Bayes' rule, compute $\mu_y(1)$ and $\sigma_y(1)$. See sections 2.5 and 2.6 for univariate and multivariate computation of the posterior.
4. Update the ensemble members. Compute the updated ensemble $\epsilon(1)$ by shifting $\tilde{\epsilon}(1)$ according to the posterior distribution of observations. See sections 2.5 and 2.6 for univariate and multivariate update rules.
5. For the assimilation of the next data point, start from pt.1 and consider $\epsilon(1)$ as your initial condition.

#### 2.5 Single observations

The case with a single vector of observations, and its errors, is the simplest and most geometrically accessible.

$$\sigma_y^2(t) = \frac{\sigma_{\tilde{y}}^2 \sigma_\omega^2}{\sigma_{\tilde{y}}^2 + \sigma_\omega^2} \tag{18}$$

$$\mu_y = \frac{\sigma_y^2}{\sigma_{\tilde{y}}^2}\mu_{\tilde{y}} + \frac{\sigma_y^2}{\sigma_\omega^2}\mu_\omega \tag{19}$$

The update formula for the j-eth observed variables in the i-eth ensemble member,

$$\vec{x}_{i,j}(t) = (\tilde{x}_i(t-1) - \mu_{\tilde{y}})\sqrt{\frac{\sigma_y^2}{\sigma_{\tilde{y}}^2}} + \mu_y, \tag{20}$$

where the squared standard deviation is kept for clarity.

And the update formula for the j-eth unobserved variable in the i-eth ensemble member is:

$$x_{i,j}(t) = \tilde{x}_{i,j}(t) + p_1(y - \tilde{y}), \tag{21}$$

where $p_1$ is such that $x = p_0 + p_1 y$ is the least squares fit, or, in this case $p_1 = \sigma_{\tilde{x}\tilde{y}}/\sigma_{\tilde{y}}^2$.

#### 2.6 Multiple observations

When using multiple observations in a time point, we need to use multivariate normal distributions. The product $\mathcal{N}_3(\vec{\mu}_3, \Sigma_3)$, of two probability density functions $\mathcal{N}_1(\vec{\mu}_1, \Sigma_1)$ and $\mathcal{N}_2(\vec{\mu}_2, \Sigma_2)$ can be computed from its individual parameters:

$$\Sigma_3 = \Sigma_1 s^{-1} \Sigma_2 \tag{22}$$

$$\vec{\mu}_3 = \Sigma_2 s^{-1} \vec{\mu}_1 + \Sigma_1 s^{-1} \vec{\mu}_2 \tag{23}$$

with $s^{-1} = (\Sigma_1 + \Sigma_2)^{-1}$, these expressions reduce the computation cost as only one matrix inverse is needed.

The update formula for the j-eth observed variable in the i-eth ensemble member is:

$$x_{i,j} = (y_{i,j} - \mu_{\tilde{y}_j}) V_1 D_1^{-1} D_2 V_2^{-1} + \mu_{y_j}, \tag{24}$$

where $V_x$ and $D_x$ are the diagonalization matrices of $\Sigma_x$ such that $\Sigma_x V_x = V_x D_x$.

An the formula for the j-eth unobserved variable in the i-eth ensemble member is:

$$x_{i,j}(t) = \tilde{x}_{i,j}(t) + \sum_{k=1}^{N} p_k(y_k - \tilde{y}_k), \tag{25}$$

where $p_k$ is such that $x = p_0 + p_1 y_1 + ... + p_N y_N$ is the least squares fit solution.

#### 2.7 Single rank correlations

Perhaps, the linear least squares fit becomes a poor fitting method, apparent when there is a non-linear relationship between the prior parameter distribution and the prior observable distribution. We can then improve its performance with non-linear fits, but this would imply prior knowledge of the shape of this relationship. When this is not available, and the relationship is monotonic, we can resort to rank correlations [12]. Briefly, we transform the j-eth state variable prior distribution $\tilde{x}_{i,j}(t)$, and the prior and posterior observations $\tilde{y}_i(t)$ and $y_i(t)$, respectively, into their corresponding ranks $\tilde{r}^x_{i,j}(t)$ and $\tilde{r}^y_i(t)$. We perform the update in generalized continuous ranks, and then revert them into variable values. Here is an example code in `matlab`:

```
% Step 1. Compute ranks of a prior state variable and prior observable.
rx = tiedrank(State_prior);
ry = tiedrank(Obs_prior);

% Step 2. Compute generalized rank of the posterior observations
ry_ = interp1(Obs_prior, ry, Obs_post,'linear','extrap');

% Step 3. Compute observation rank increments
Dry = (ry_-ry);

% Step 4. Compute the regression between rank statistics
fr = fit(rx,ry,'poly1');

% Step 5. Compute posterior state generalized rank
rx_ = rx + fr.p1.*Dry;

% Step 6. Convert the state posterior ranks, into state posterior values
State_post = interp1(rx,State_prior,rx_,'linear','extrap');
```

#### 2.8 Inflation avoids divergence

It is common to find that the bare algorithm itself tends to reduce the ensemble variance, and although this is desirable to some extent, when the ensemble members are almost identical to each other it may cause model divergence, which leads to assimilation problems [10, 11]. The most relevant to us was that since the prior distribution becomes extremely narrow compared to observational variance,i.e. $\sigma^2_\omega \ll \sigma^2_{\tilde{y}}$, then equations 18 and 19 yield a posterior distribution essentially equal to the prior, in other words, observations are ignored. To compensate for this "excessive speed" in reducing the prior variance, we can add an inflation factor to all,

$$x_{i,j} \leftarrow \sqrt{\lambda}(x_{i,j} - \langle x_j \rangle_i) + \langle x_j \rangle_i, \tag{26}$$

where $\langle x_j \rangle_i = \sum_{i=1}^{S} x_{i,j}/S$ is the mean value of the j-eth variable across all ensemble members. Typical values for inflation are $\lambda = 1.02, 1.05$ or even $1.10$.

There is also the possibility of adding a time-dependent inflation that, for example depends on the distance between your likelihood and prior distributions.

### References

1. Bjørnstad, O. N., Shea, K., Krzywinski, M. & Altman, N. The SEIRS model for infectious disease dynamics. *Nature Methods* **17,** 557–558. ISSN: 1548-7105. https://doi.org/10.1038/s41592-020-0856-2 (June 2020).
2. Rees, E. M. *et al.* COVID-19 length of hospital stay: a systematic review and data synthesis. en. *medRxiv.* Publisher: Cold Spring Harbor Laboratory Press, 2020.04.30.20084780. https://www.medrxiv.org/content/10.1101/2020.04.30.20084780v3 (2020) (May 2020).
3. Armstrong, R. A., Kane, A. D. & Cook, T. M. Outcomes from intensive care in patients with COVID-19: a systematic review and meta-analysis of observational studies. en. *Anaesthesia,* anae.15201. ISSN: 0003-2409, 1365-2044. https://onlinelibrary.wiley.com/doi/abs/10.1111/anae.15201 (2020) (July 2020).
4. *ICNARC – Reports* https://www.icnarc.org/Our-Audit/Audits/Cmp/Reports (2020).
5. *Report of the WHO-China Joint Mission on Coronavirus Disease 2019 (COVID-19)* en. Library Catalog: www.who.int. https://www.who.int/publications-detail-redirect/report-of-the-who-china-joint-mission-on-coronavirus-disease-2019-(covid-19) (2020).
6. *The Epidemiological Characteristics of an Outbreak of 2019 Novel Coronavirus Diseases (COVID-19) — China, 2020* http://weekly.chinacdc.cn/en/article/doi/10.46234/ccdcw2020.032 (2020).
7. Bar-On, Y. M., Flamholz, A., Phillips, R. & Milo, R. SARS-CoV-2 (COVID-19) by the numbers. *eLife* **9** (ed Eisen, M. B.) Publisher: eLife Sciences Publications, Ltd, e57309. ISSN: 2050-084X. https://doi.org/10.7554/eLife.57309 (2020) (Mar. 2020).
8. Ma, J. Estimating epidemic exponential growth rate and basic reproduction number. *Infectious Disease Modelling* **5,** 129–141. ISSN: 2468-0427. http://www.sciencedirect.com/science/article/pii/S2468042719300491 (2020).
9. Diekmann, O., Heesterbeek, J. A. P. & Roberts, M. G. The construction of next-generation matrices for compartmental epidemic models. eng. *Journal of the Royal Society, Interface* **7,** 873–885. ISSN: 1742-5662. https://doi.org/10.1098/rsif.2009.0386 (June 2010).
10. Evensen, G. *Data assimilation: the ensemble Kalman filter* 2nd ed. ISBN: 978-3-642-03710-8 (Springer, Dordrecht ; New York, 2009).
11. Asch, M., Bocquet, M. & Nodet, M. *Data assimilation: methods, algorithms, and applications Fundamentals of algorithms* **11.** ISBN: 978-1-61197-453-9 (SIAM, Society for Industrial and Applied Mathematics, Philadelphia, 2016).
12. Anderson, J. L. A Nonlinear Rank Regression Method for Ensemble Kalman Filter Data Assimilation. en. *Monthly Weather Review* **147,** 2847–2860. ISSN: 0027-0644, 1520-0493. https://journals.ametsoc.org/mwr/article/147/8/2847/344624/A-Nonlinear-Rank-Regression-Method-for-Ensemble (2020) (Aug. 2019).

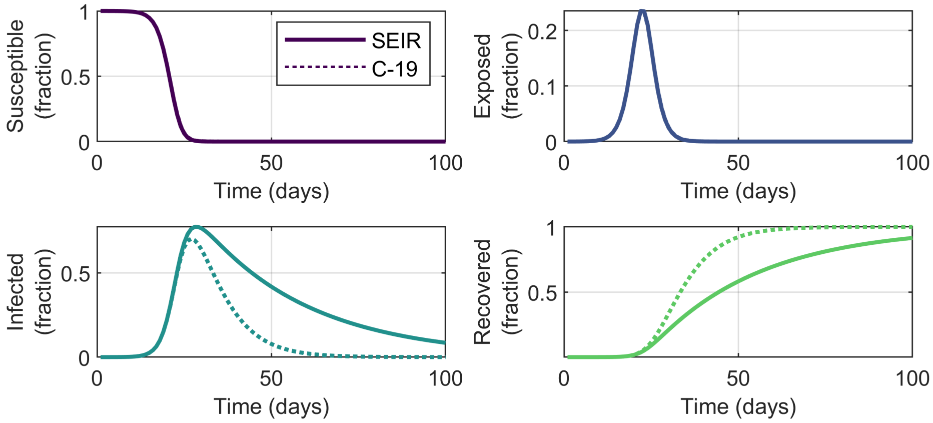

**Figure S1. A minimal** `SEIRS` **model is not completely equivalent to our COVID-19 model.** We obtained an equivalence between the specific parameters of our COVID-19 and a minimal `SEIRS` model, to find that the dynamics of our COVID-19 model cannot be fully captured by the simpler `SEIRS` model. Observe however the good agreement between both models during the initial days.

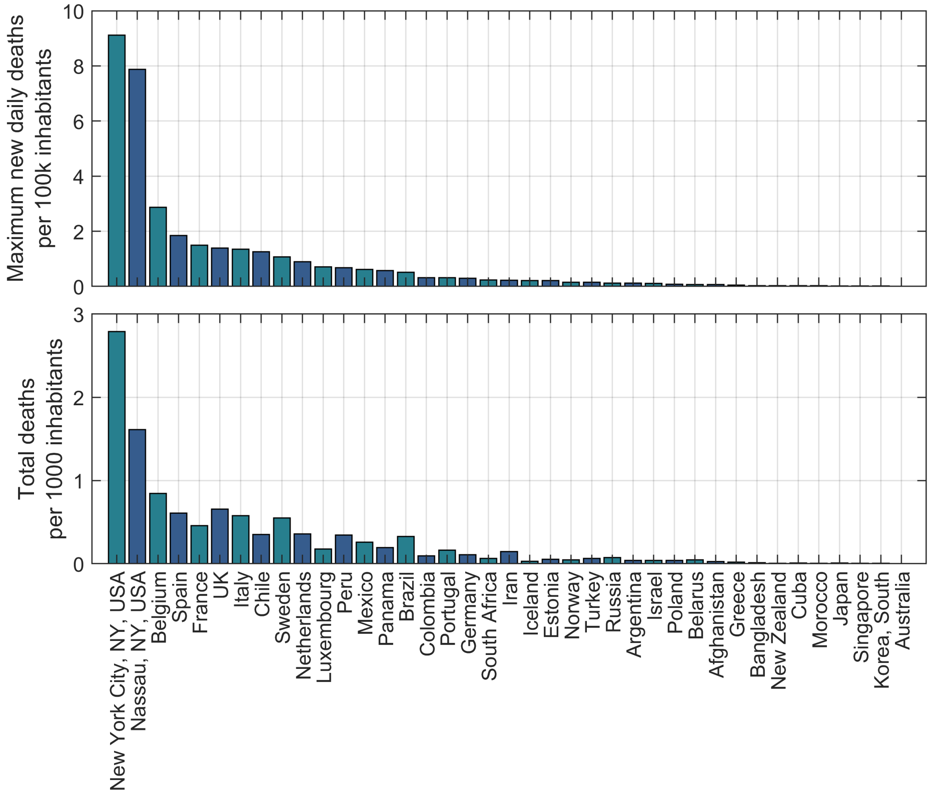

**Figure S2. Impact of COVID-19 upon different countries and regions.** We assessed the impact of the COVID-19 epidemic upon 38 countries and 2 counties in the USA by measuring (Top) the maximum daily deceases per $10^5$ inhabitants and (Bottom) the total deceases per $10^3$ inhabitants. Two USA counties together with Belgium, Spain and France are the most promising candidates. However, we discarded data from Nassau as it was considerably less smooth than the rest.

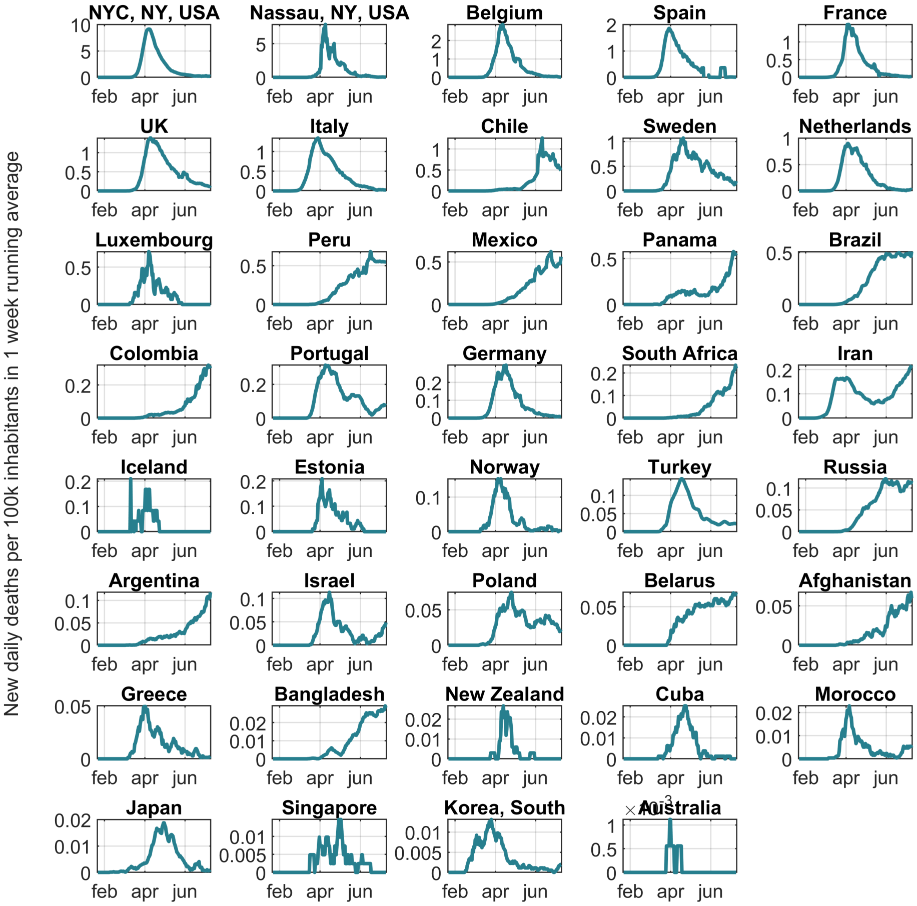

**Figure S3. Daily deceases of COVID-19 upon different countries and regions.** We show the 14 day running average of COVID-19 daily deceases reported in different regions. Data is normalized with respect to the total population of the corresponding region, which indicates the severity of the impact of COVID-19.

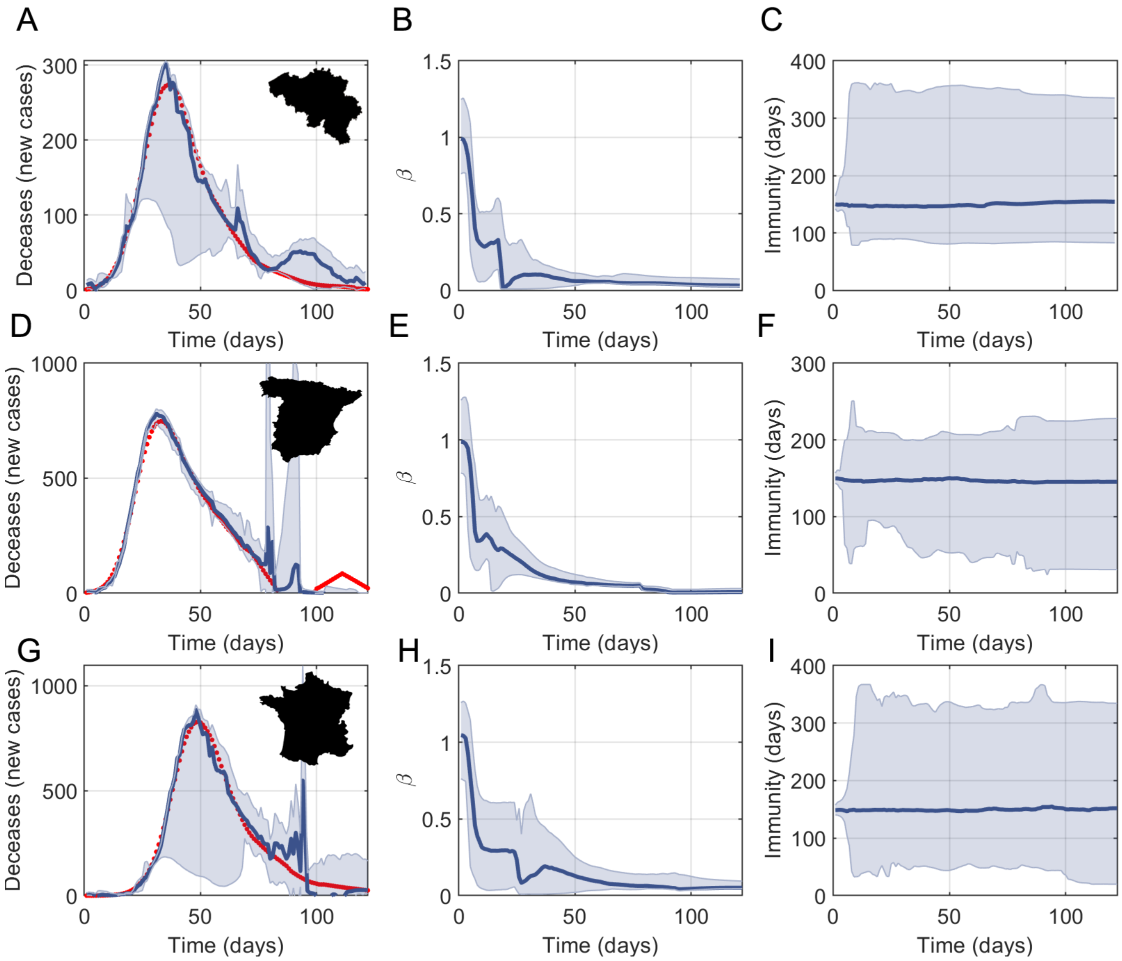

**Figure S4. EAKF results for Belgium, Spain and France.** Apart from New York City, we tested our approach with data of three other heavily-affected countries: (A-C) Belgium, (D-F) Spain and (G-I) France. (A,D and G) Data (red dots) and protocol estimate (median and 95% CI in blue) of daily deceases. (B,E and H) Model estimates of the infection rate in days$^{-1}$. (C,F and I) Protocol estimates of the immune memory duration $\tau$.

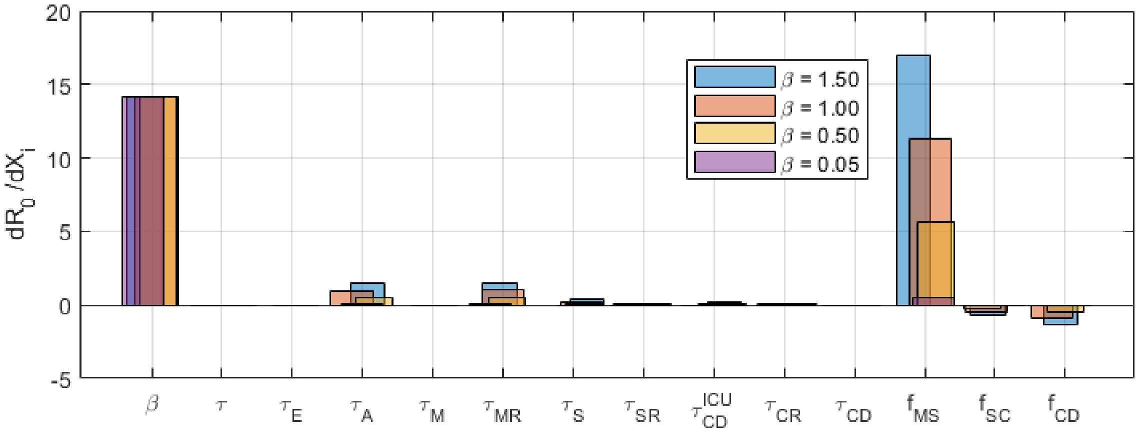

**Figure S5. Parameter sensitivity of $R_0$.** We computed the numerical value of the local partial derivative of $R_0$ with respect to every other model parameter. We find that $R_0$ strongly depends, in all cases, on $\beta$. However it also depends strongly on $f_{MS}$, the fraction of mild cases that worsen, which supports the importance of prevention, early detection of symptoms and palliative care measures.
